# Determining Personalized Community Health Needs by Feature Selection and Clustering

**DOI:** 10.1101/2020.02.21.20024612

**Authors:** Matthew Agar-Johnson

**Affiliations:** Dept of Computational Biology, Carnegie Mellon University, Pittsburgh, PA

## Abstract

The Center for Disease Control, through the Community Health Data Initiative (CHDI), has released a large dataset by county detailing the overall health indicators, demographics, and major risk factors and causes of morbidity and mortality in the US. In order to address the heterogeneity of community healthcare in the US, k-Means clustering was performed on the CHDI dataset to determine community subtypes in terms of health challenges and outcomes. The optimal number of eight clusters was determined by the Elbow Method, and clusters were analyzed to determine significant differences in demographic. In order to determine community-specific healthcare solutions and directions, feature selection and modeling of healthcare outcomes was performed for each of the eight subtypes using LASSO regression. It was determined that different features significantly impact health outcomes in the different clusters, providing information about the unique health challenges faced by these different types of communities. LASSO regression using the entire unclustered dataset yielded significantly poorer results on the sub-clusters in terms of model performance, further supporting the claim that modeling community-specific needs is a vital step for delivering accurate and adequate community healthcare. These results have the potential to inform policymaking at the local/municipal level, as well as inform the approaches taken by primary practitioners to address community needs.

## Introduction

The Center for Disease Control, through the Community Health Data Initiative (CHDI), has released a large dataset by county detailing the overall health indicators, demographics, and major risk factors and causes of morbidity and mortality in the US[1]. In order to address the heterogeneity of community healthcare in the US, k-Means clustering was performed on the CHDI dataset to determine community subtypes in terms of health challenges and outcomes. The optimal number of eight clusters was determined by the Elbow Method, and clusters were analyzed to determine significant differences in demographic. In order to determine community-specific healthcare solutions and directions, feature selection and modeling of healthcare outcomes was performed for each of the eight subtypes using LASSO regression. It was determined that different features significantly impact health outcomes in the different clusters, providing information about the unique health challenges faced by these different types of communities. LASSO regression using the entire unclustered dataset yielded significantly poorer results on the sub-clusters in terms of model performance, further supporting the claim that modeling community-specific needs is a vital step for delivering accurate and adequate community healthcare. These results have the potential to inform policymaking at the local/municipal level, as well as inform the approaches taken by primary practitioners to address their communitys needs.

## Background

### Data

The CHDI data were released with the intention of encourage[ing] dialogue about actions that can be taken to improve community health, and presentindicators like deaths due to heart disease and cancer[1]. The data are presented by county in the United States (3142 counties) and contain a variety of demographic and health outcome features, such as racial, age, and gender demographics, income demographics, access to healthcare, and health outcomes. In recent years, cluster analysis has begun to be applied to large health-care datasets with the aim of determining specific subgroups of individual patients and predicting health outcomes, but has not yet been applied to healthcare at the community level[2].

### Clustering by k-Means

k-Means is a clustering method generally accepted for determining significant clusters in large datasets in an unsupervised manner. It is a centroid based, iterative approach that requires a predefined number of clusters k. The algorithm begins by defining a set number k of cluster centers {*c*_1_, *c*_2_, *c*_*k*_}. Each of the k clusters is defined by its center, or centroid. The first step is the assignment step, where each point in the dataset is assigned to the cluster with the nearest center in terms of Euclidean distance. In the second step, the update step, Each of the centroids is re-defined as the mean of all datapoints assigned to that cluster. These two steps proceed iteratively until convergence [3].

There exist several methods to determine the optimal numbers of clusters that vary in terms of complexity. The most nave approach is the elbow method, where the data is clustered with varying number of clusters k and the sum of squared-errors (SSE) within is cluster is computed[4]. Normally, as the number of clusters increases, the within-cluster SSE decreases sharply at first, then inflects and begins decreasing slowly. This inflection point, or elbow, is the region from which to select the optimal value for k.

### Feature Selection by LASSO Regression

Once the clusters have been determined, the next step is to determine relevant features for each community subtype. This is generally known as a feature selection problem, and there exist a variety of methods for solving these problems, both for binary/multinomial classification problems and for continuous class variables. Least absolute shrinkage and selection operator (LASSO) regression is a variation of continuous variable feature-selection based on linear regression. It introduces an L1 regularization penalty, meaning minimizing the objective function:

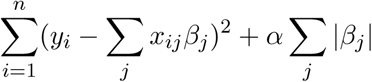

Where *β*_*j*_ are feature coefficients and *α* is a scaling parameter indicating the strength of the L1 penalty[5]. In other words, there is a penalty introduced that corresponds directly with the number of features selected, i.e. the cardinality of the non-zero coefficient vector. In minimizing this objective function, the regressor will learn the best possible model with the fewest number of features, depending on the magnitude of the parameter. The higher the value of this parameter, the fewer features will be selected, at the cost of overall fit of the model. For the problem examined in this study, *α* should be tuned such that a small, interpretable number of features are selected without sacrificing too much in terms of model fit.

## Materials and Methods

### Data Pre-Processing

When the clustering algorithm is run, there are several features in the dataset that may have significant influence over the resulting clustering. This include features that may have significantly higher variance than others, or features that strongly correlate with others in the data. The most notable example of this is population size, which differs vastly between large urban centers (Cook County, IL and Los Angeles, CA are two examples), and smaller rural communities with low population density. Other features, such as absolute numbers of uninsured and elderly individuals, also vary wildly as they correlate directly with population. To control for this, and ensure that these factors did not overwhelm the clustering, the population feature was removed from the feature space, and any features based on absolute numbers of individuals were converted to their respective per capita numbers.

Before performing clustering or feature selection, data normalization is required to account for different scales between features. The method chosen for this study was min-max normalization, which converts each feature value according to:

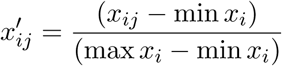

The resulting normalized data consists of 3142 observations of 41 features, and can then be clustered by the k-Means algorithm.

### Cluster Optimization

Using the scikit-learn k-Means package, the pre-processed data was cluster with 1k30, and for each resulting clustering, the intra-cluster SSE was evaluated. This data was then plotted and observed for a single inflection point. The inflection point indicates the optimal number of clusters for subsequent steps.

### Clustering by k-Means

Once the optimal number of clusters k had been determined, the data was clustered using k-Means. Each cluster was then analyzed by the mean min-maxed value for each feature, and the values for each cluster were plotted to determine community differences.

### Feature Selection and LASSO Regression

When considering feature selection in this problem of community health outcomes, it is important to consider a number of facets of the dataset itself. In our case, health outcomes, regardless of community subtype, will be most significantly affected by features that are guaranteed to be common indicators of health outcomes across all communities. These include rate of smoking, obesity, and diabetes, as well as features that correlate with poverty and access to healthcare. Nave feature selection may not reveal significantly pertinent information if there is not control for these overwhelming features. To this effect, smoking, obesity, and diabetes were removed from the feature space.

The L1 penalty parameter *α* was tuned by plotting the mean *R*^2^ correlation against the value of alpha for varying levels of the parameter, as well as the cardinality of the non-zero selected feature set. The selection of the parameter was somewhat arbitrary, and was determined by a value that resulted in decent model performance (*R*^2^ *∼* 0.6 for each cluster), while maintaining a small enough feature set to be easily interpretable. All model performance analysis was performed using a random 80/20 train/test split.

Once the parameters of the regression model had been tuned, each cluster was analyzed by LASSO regression. Regression was also performed on the full unclustered dataset, to produce a set of non community-specific features that could be used to compare model performance against models trained on the clustered data. The feature sets for each cluster were stored in a table, and *R*^2^ scores were computed for each cluster using both the clustered feature set and using the feature set generated by the unclustered data. This comparison is intended to emphasize the specific and different health challenges of different communities and to validate the significance of our approach.

## Results

### Cluster Optimization

As seen in Figure 1, there is a single inflection point observed between 7 and 8 clusters. The elbow method is a somewhat nave method, and the differences in model performance between 7 and 8 clusters is not determinable absolutely using this method. A final cluster number of 8 was selected due to there being greater consistency in the size of clusters for this number than for k=7.

**Figure 1:**
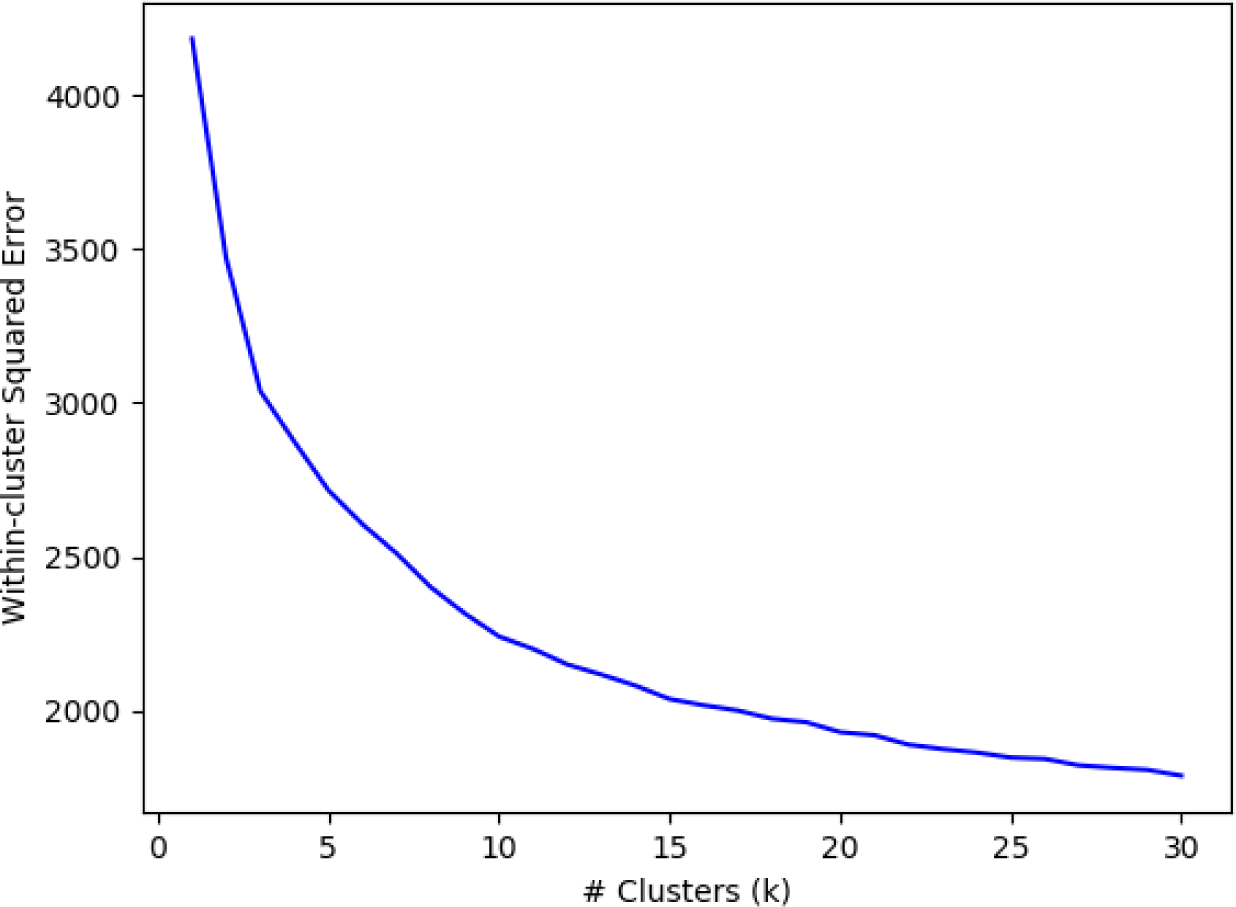
Within-cluster sum of squared errors (SSE) against varying number of clusters k for unsupervised clustering of the CHDI dataset. A single inflection point is observed between 7 and 8 clusters, indicating the optimal value for k. Clustering was performed using the sklearn k-Means package.

### Cluster Analysis by k-Means

Overall, significant demographic heterogeneity was observed between clusters. Cluster 1 showed high levels of population density and Asian population, as well as high access to healthcare, high insurance rates, and an overall higher-than-average health status. Cluster 1 skews toward California, the Northeastern US, and the Mid-Atlantic region (Pennsylvia, Maryland). Cluster 2 contains higher incomes, greater access to healthcare, above-average white population, and overall good health. Cluster 2 skews toward the Mid- and North-west, as well as a number of counties in Texas and along the East and West coasts of the US. Cluster 3 shows high poverty and obesity, skews toward Black and Hispanic population, has low access to healthcare, and shows overall lower health status. Cluster 3 skews overwhelmingly toward the southeastern US (Alabama, Georgia, Mississippi, Louisiana). Cluster 7 shows low population density and insurance rates, lower use of healthcare, and poorer overall health outcomes. Cluster 7 contains low population-density counties primarily from Appalachian states and central states (Indiana, North Dakota, Montana, Idaho).

### LASSO Regression Parameter Tuning

The mean number of features selected as well as overall average *R*^2^ score for 100 rounds of randomized 80/20 cross-validation were performed and plotted against varying values of the L1 penalty parameter *α*.

The value of *α* = 0.2 was observed to have a sharp decrease in the cardinality of the feature set while maintaining model performance. An average number of features equal to 6 was considered optimal for interpretation. Notably, even with this low-magnitude L1 penalty, in Cluster 2 only two features were selected, so this parameter was set to 0.2 for subsequent steps so that some interpretable data from all clusters could be determined.

### Feature Selection by LASSO and Analysis

LASSO regression was performed against the overall health-status indicator feature Health Status, first on the entire unclustered dataset, then on each cluster independently.

The features are presented in Figure 4 in order of the absolute value of coefficients.

**Figure 2:**
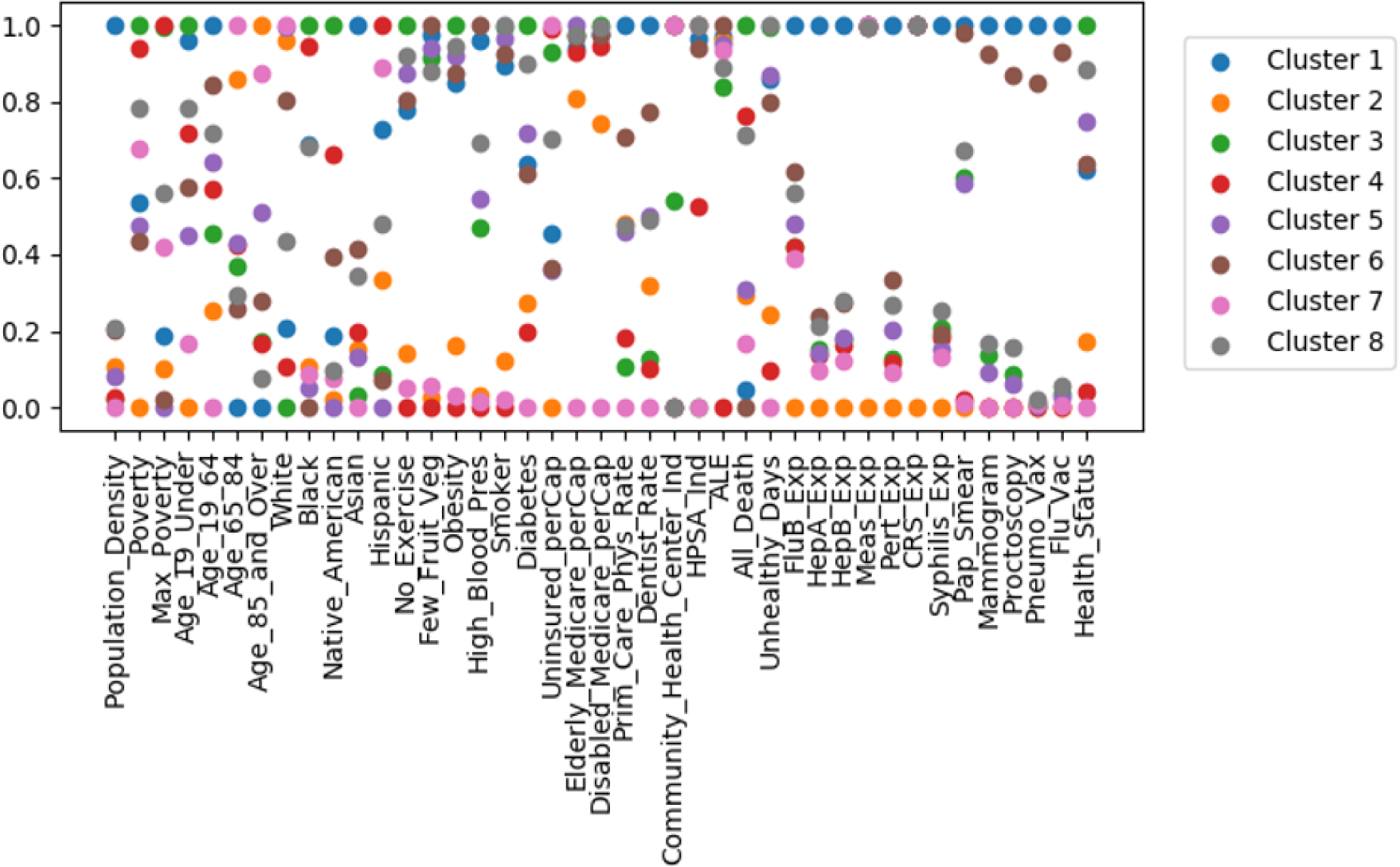
Mean per-cluster feature values following clustering of CHDI dataset by k-Means. Feature values were normalized using min-max normalization and the mean values of each feature for every cluster are computed and plotted. A value of 1.0 indicates a values equal to the max among all datapoints, and a value of 0.0 indicates a value equal to the min among all datapoints

**Figure 3:**
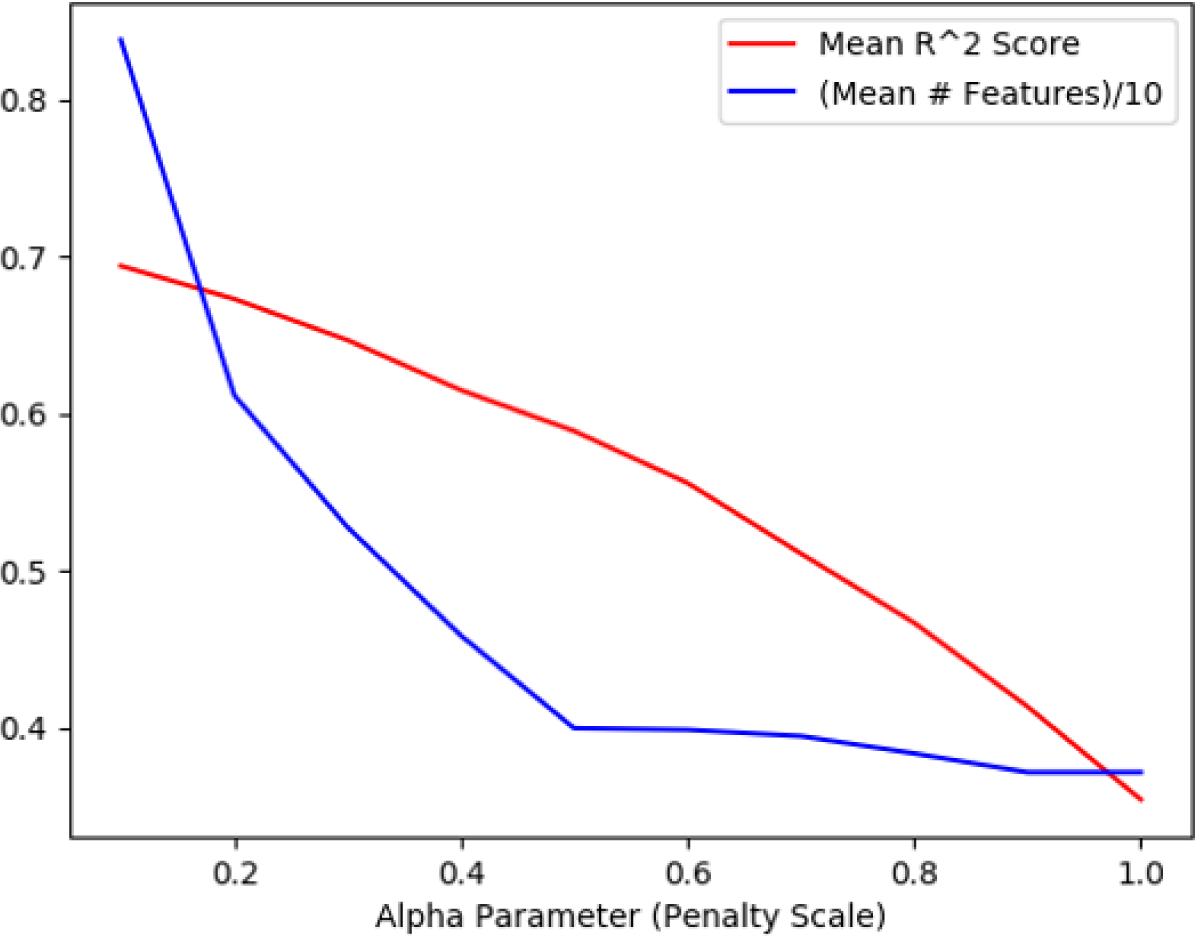
Mean *R*^2^ score (red) and 0.1*(mean num. selected features) (blue) for varying levels of L1 parameter *α*. Scores were computed by 100-fold 80/20 cross-validation. The level of 0.6 for the blue line at *α* = 0.2 represents a mean number of 6 features selected by LASSO regression.

**Figure 4:**
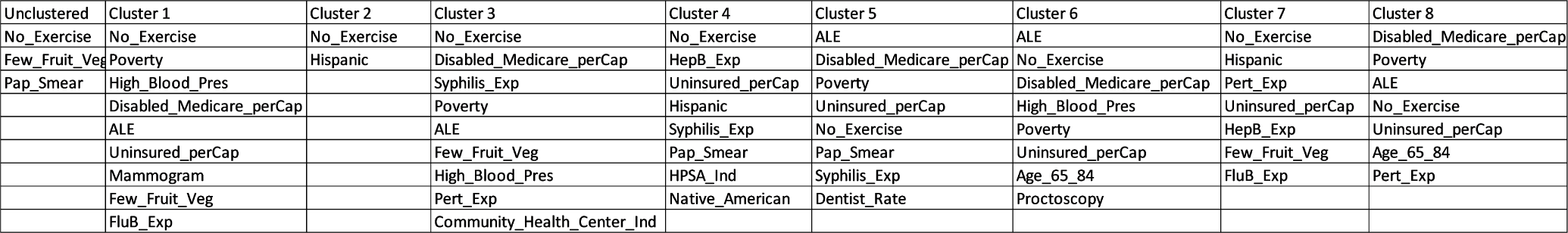
Selected Features using LASSO regression (*α* = 0.2), for unclustered CHDI data and for eight clustered determined previously by k-means. Features are presented, from top to bottom, in order of the highest absolute-value coefficients. Regression was performed against the Health Status feature.

To determine the significance of the difference observed in feature sets between clusters, the regression was performed again, first using the features selected for each cluster, then using the features selected for the unclustered data. The results are shown in Figure 5.

**Figure 5:**
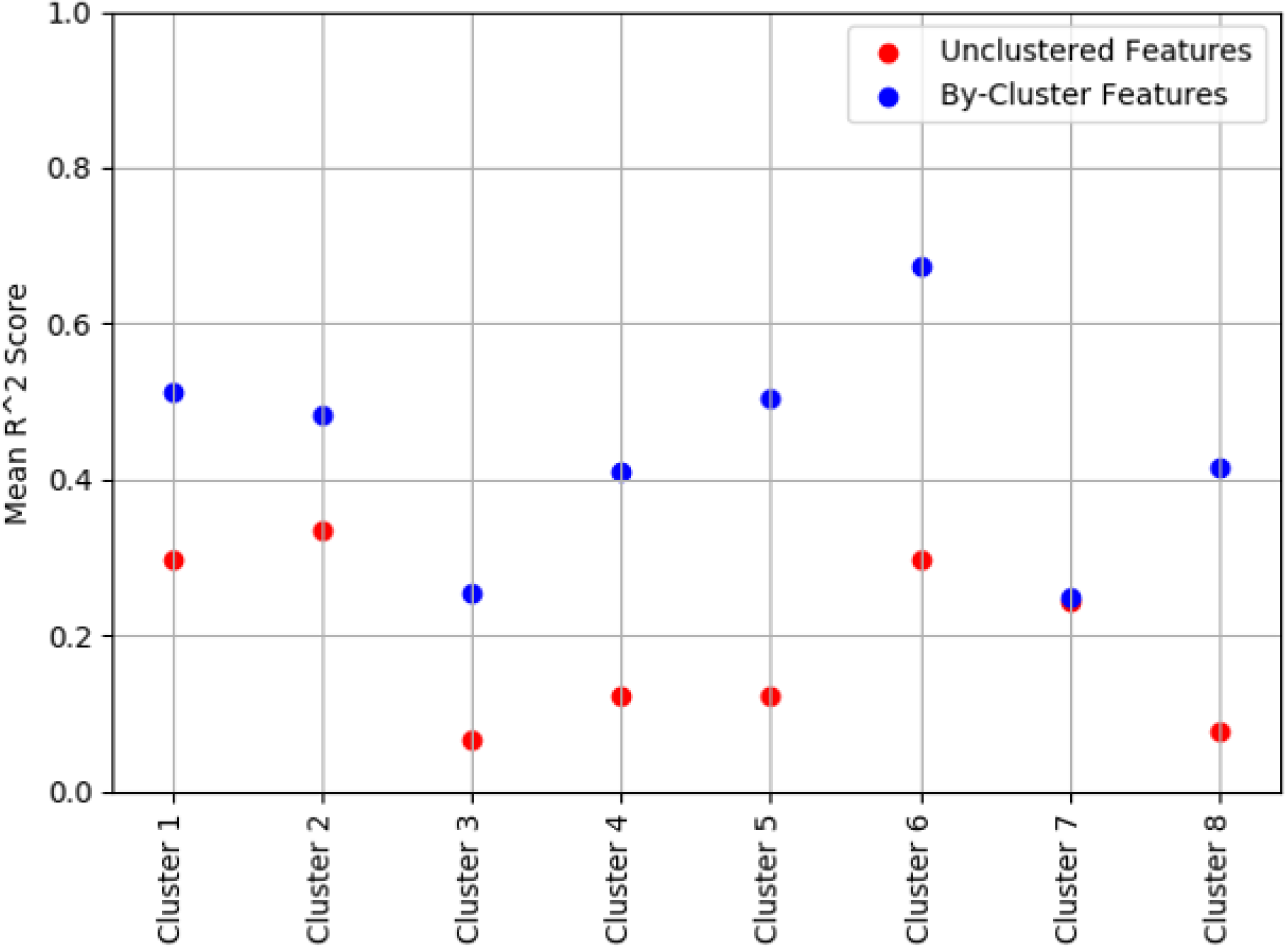
Average *R*^2^ scores for LASSO regression on unclustered CHDI data and all clusters using the features selected for unclustered data (red) and using features obtained from LASSO on each cluster individually (blue). Scores were computed using 100-fold 80/20 split cross-validation with random sampling. *p* < 0.01 for clusters 1-6 and cluster 8, and *p* < 0.05 for cluster 7.

Significant increases in model performance (*p* < 0.01) are observed in all clusters except Cluster 7 when using cluster-specific features compared to the unclustered, and *p* < 0.05 is observed for cluster 7.

## Discussion

### Clustering

The results of the k-means clustering were not too surprising considering a priori knowledge about the demographic and regional differences in the United States, but the appearance of trends by easily interpretable demographics indicates that the method used in this study was reasonably robust. The conclusions obtained from unsupervised clustering of the data support intuitions about demographic and regional differences, while counties that appear distal from geographic centers within a cluster suggest that geography and population density alone are not fully indicative of which community-type a given country may be (full list of counties by cluster included in supplement). Overall trends of mortality and morbidity by cluster may provide insight to health policymakers and physicians alike in terms of which communities are lacking healthcare access and beneficial outcomes, and may require the most attention and resources.

### Feature Selection/LASSO

Figure 4., the table showing a list of features selected for modeling health status in each cluster, reveal some surprising, and perhaps informative, results. In clusters 2, 4, and 7, belonging to the Hispanic demographic has a significant effect on overall health status. This may indicate that in certain types of communities, the healthcare needs of Hispanic communities are not being properly addressed. The same can be said for the selection of the Native American demographic in Cluster 4. The clustering also reveals that different types of communities may have different diseases that contribute to overall negative health outcomes. Influenza B appears to be a significant effector of health outcomes in clusters 1 and 7, Pertussis appears in clusters 3, 7 and 8, Syphilis in clusters 4 and 5, and Hepatitis B in clusters 4 and 7. Since many of these diseases are vaccine-preventable, there may be significant health improvements to be made in these community-types by more effective vaccination programs. In clusters 6 and 8, elderly populations are seen to have a large effect on health outcomes, suggesting possible improvements in elderly care in these community types. Importantly, none of these features are prominent when the data is modeled in an unclustered manner.

The importance of the clustering method described in this study is further supported by Figure 5., where significant improvements in model performance over the clustered data were observed in all clusters when feature selection was performed independently on each cluster rather than on the dataset as a whole. Many of the values observed in figure 5 are lower than the 0.6 value observed in Figure 3, due to the omission of the features corresponding to obesity, smoking and diabetes, which were omitted due to their high predictive power across all clusters. The results in Figure 5 not only support the hypothesis of heterogeneous nature of community health needs across demographics, but further demonstrate the robustness of our approach.

## Conclusions

All of these computational methods are ultimately a tool to support primary care practitioners and health policymakers in individualizing and personalizing healthcare at the community level. Due to the limited knowledge of community/individual medical health by the authors of the study, logical next steps would be the collaboration with health policymakers and physicians to confirm the validity of the findings, and to suggest further directions for more targeted evaluation of these demographic and healthcare markers in different sub-populations.

## Data Availability

All data is publicly available from the CDC website at https://healthdata.gov/dataset/community-health-status-indicators-chsi-combat-obesity-heart-disease-and-cancer

## Notes

### Competing Interest Statement

This study is published independently and does not carry the endorsement of Carnegie Mellon University.

### Funding Statement

No funding was used for this study.

